# The Clinical Value of ChatGPT for Epilepsy Presurgical Decision Making: Systematic Evaluation on Seizure Semiology Interpretation

**DOI:** 10.1101/2024.04.13.24305773

**Authors:** Yaxi Luo, Meng Jiao, Neel Fotedar, Jun-En Ding, Ioannis Karakis, Vikram R. Rao, Melissa Asmar, Xiaochen Xian, Orwa Aboud, Yuxin Wen, Jack J. Lin, Fang-Ming Hung, Hai Sun, Felix Rosenow, Feng Liu

## Abstract

**Background:** For patients with drug-resistant focal epilepsy (DRE), surgical resection of the epileptogenic zone (EZ) is an effective treatment to control seizures. Accurate localization of the EZ is crucial and is typically achieved through comprehensive presurgical approaches such as seizure semiology interpretation, electroencephalography (EEG), magnetic resonance imaging (MRI), and intracranial EEG (iEEG). However, interpreting seizure semiology poses challenges because it relies heavily on expert knowledge and is often based on inconsistent and incoherent descriptions, leading to variability and potential limitations in presurgical evaluation. To overcome these challenges, advanced technologies like large language models (LLMs)—with ChatGPT being a notable example—offer valuable tools for analyzing complex textual information, making them well-suited to interpret detailed seizure semiology descriptions and assist in accurately localizing the EZ.

**Objective:** This study evaluates the clinical value of ChatGPT in interpreting seizure semiology to localize EZs in presurgical assessments for patients with focal epilepsy and compares its performance with epileptologists.

**Methods:** Two data cohorts were compiled: a publicly sourced cohort consisting of 852 semiology-EZ pairs from 193 peer-reviewed journal publications and a private cohort of 184 semiology-EZ pairs collected from Far Eastern Memorial Hospital (FEMH) in Taiwan. ChatGPT was evaluated to predict the most likely EZ locations using two prompt methods: zero-shot prompting (ZSP) and few-shot prompting (FSP). To compare ChatGPT’s performance, eight epileptologists were recruited to participate in an online survey to interpret 100 randomly selected semiology records. The responses from ChatGPT and the epileptologists were compared using three metrics: regional sensitivity (RSens), weighted sensitivity (WSens), and net positive inference rate (NPIR).

**Results:** In the publicly sourced cohort, ChatGPT demonstrated high RSens reliability, achieving 80-90% for the frontal and temporal lobes, 20-40% for the parietal lobe, occipital lobe, and insular cortex, and only 3% for the cingulate cortex. The WSens, which accounts for biased data distribution, consistently exceeded 67%, while the mean NPIR remained around 0. These evaluation results based on the private FEMH cohort are consistent with those from the publicly sourced cohort. A group *t*-test with 1000 bootstrap samples revealed that ChatGPT-4 significantly outperformed epileptologists in RSens for commonly represented EZs, such as the frontal and temporal lobes (p < 0.001). Additionally, ChatGPT-4 demonstrated superior overall performance in WSens (p < 0.001). However, no significant differences were observed between ChatGPT and the epileptologists in NPIR, highlighting comparable performance in this metric.

**Conclusions:** ChatGPT demonstrated clinical value as a tool to assist the decision-making in the epilepsy preoperative workup. With ongoing advancements in LLMs, it is anticipated that the reliability and accuracy of LLMs will continue to improve in the future.

## Introduction

Epilepsy is one of the most common neurological diseases affecting more than 70 million people worldwide [1], with approximately 50.4 per 100,000 people developing new-onset epilepsy each year [2, 3]. For patients with drug-resistant focal epilepsy (DRE), surgical resection of the epileptogenic zone (EZ) provides an effective means to control seizure attacks.

EZs is a theoretical definition given by Dr. Hans Lüders [8] and whose removal will make the patients seizure-free, thus it can only be derived or validated after surgical oblation. Seizure semiology, which refers to signs and symptoms exhibited and experienced by a patient during epileptic seizures, yields valuable clues on localizing the EZs [5, 6]. In addition to semiology, presurgical evaluation involves multimodal brain imaging tools such as electroencephalography (EEG), stereo-electroencephalography (SEEG), magnetoencephalography (MEG), magnetic resonance imaging (MRI), and functional MRI (fMRI) [7, 8]. To determine the ground truth of EZs, the post-surgical outcome information (whether or not the patient achieved seizure freedom) is used to validate the resected brain regions, with seizure-free status determined according to the International League Against Epilepsy (ILAE) criteria “Class I: Completely seizure-free; no auras” [9] or Engel’s classification “Class I: Seizure free or no more than a few early, non-disabling seizures” [10].

Interpreting seizure semiology is a nontrivial task because it relies heavily on expert knowledge and is often based on inconsistent and incoherent descriptions, leading to variability and potential limitations in presurgical evaluation. A recent study employed the conditional inference tree algorithm to interpret seizure semiology but achieved a maximum accuracy of only 56.1% across five ictal onset regions [11]. To address the challenges associated with interpreting semiology for the localization of EZs, advanced technologies such as large language models (LLMs) have emerged as potential solutions. LLMs, particularly ChatGPT, have demonstrated remarkable capabilities across a wide range of natural language processing (NLP) tasks, including processing complex textual descriptions. Given the descriptive nature of seizure semiology, LLMs are well-positioned to address this challenge.

ChatGPT, developed by OpenAI [12], has exhibited exceptional capability in processing and interpreting extensive textual data, by training on extensive textual datasets using supervised learning and reinforcement learning from human feedback, making it a promising tool for clinical applications, such as information retrieval, clinical decision support, and medical report generation [13, 14, 15]. Notably, A study conducted in February 2023 reported that ChatGPT successfully passed the United States Medical Licensing Examination (USMLE) [16], confirming its potential as a reliable source of medical information. The increasing application of ChatGPT in diagnosing various diseases inspired us to utilize it to interpret seizure semiology and to localize the EZs, potentially to be used as an AI co-pilot for the presurgical evaluation of patients with epilepsy [17, 18].

It is desirable to test the clinical value of LLMs, such as ChatGPT, on interpreting semiology to predict EZ localization. Given that EZs can be categorized into six distinct brain regions, it is important to further explore whether ChatGPT demonstrates varied precisions in localizing these zones. We used the LCN-CortLobes classification system (FreeSurferWiki, LCN-CortLobes) [4] to provide a standardized anatomical framework to categorize EZs into six regions: the frontal lobe, temporal lobe, parietal lobe, occipital lobe, cingulate cortex, and insular cortex. If ChatGPT achieves superior performance, particularly in specific brain regions or consistently across all regions, it would showcase its potential to assist in epilepsy presurgical decision-making.

## Methods

### IRB Approval

The Institutional Review Board (IRB) at Stevens Institute of Technology exempted the approval of this secondary data analysis study under protocol 2024-039 (N).

### Public and Private Data Cohorts

The performance of ChatGPT was first evaluated using a seizure semiology database compiled from publicly available studies in published peer-reviewed journals. Furthermore, to mitigate the risk of testing the performance with the data that may have been utilized during ChatGPT’s training, another separate private data cohort was constructed from electronic health records (EHR) at Far Eastern Memorial Hospital (FEMH) hospital in Taiwan for external validation.

The public data cohort was compiled from peer-reviewed research articles identified through a systematic search in PubMed using keywords including “seizure”, “seizures semiology”, “epilepsy”, and “epileptogenic zones” in the past 20 years [19]. Each research paper includes a combination of individual and group case reports, as well as details on the EZs. Relevant information, such as seizure semiology and patient-specific details (e.g., age, gender, and handedness), was extracted from the text or tables within the publications. All the extracted cases have both descriptions of seizure semiology and validated EZs. The EZ was validated with good surgical outcomes by the authors [20]. In cases where multiple EZs were identified, multiple general regions were assigned accordingly.

As illustrated in the top-left corner of Figure 1A, the public data cohort initially included 309 publications. Among these, 116 studies were excluded due to presenting uncertain EZs, such as those specifying only hemisphere-level EZs (e.g., “right hemispherectomy” or “left subtotal hemispherectomy”). From the remaining 193 studies, 893 cases were extracted with descriptions of seizure semiology, with additional 43 of them being excluded due to unclear or non-specific semiology descriptions, such as the vague terms (e.g., “non-specific aura”), descriptions shorter than two words, or aggregated data from large patient cohorts. Ultimately, the resulting database comprised 852 cases with detailed semiology descriptions and validated EZs. Examples of minimal semiology–EZ pairs include “speech arrest, lower of speech intensity – Insular Cortex (INS)” and “Aura with fear, aura with déjà vu – Cingulate Cortex (CING)”.

**Figure 1.**
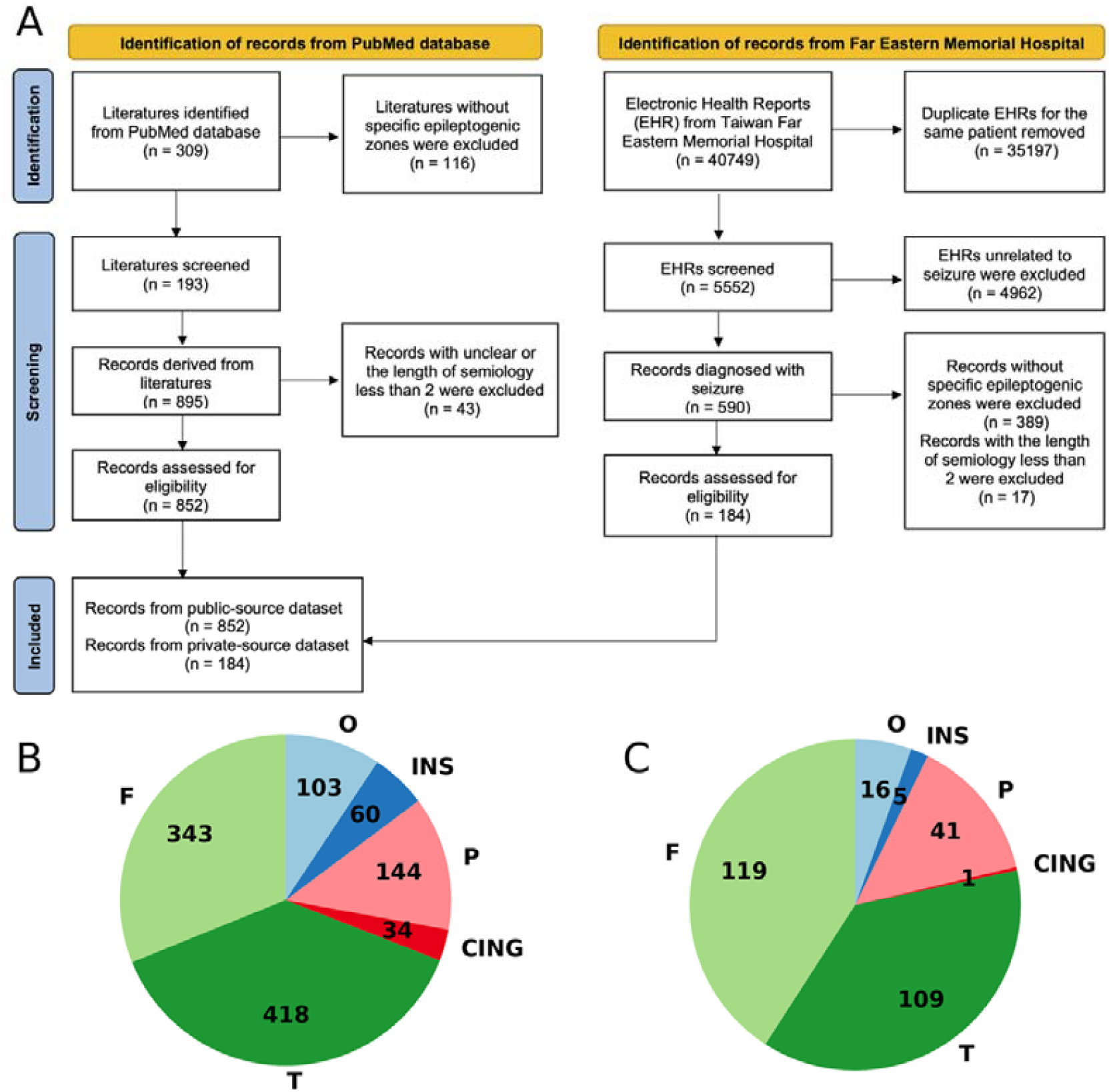
(A) PRISMA Flow Diagram for Publicly Sourced and Private-Source Database Construction. (B) Region Distribution for Publicly sourced Cohort. (C) Region Distribution for Private-Source Cohort.

The private data cohort was compiled from EHR at FEMH in Taiwan, covering the period from 2017 to 2021. These EHRs included patients’ demographic information (e.g., age, gender), clinical details (e.g., diagnosis IDs and dates), symptoms, laboratory results, and clinicians’ diagnoses. Semiology was extracted from clinicians’ notes documenting patients’ symptoms, such as “dizziness and gait disturbance since late August 2016, no dysarthria or fever.” EZs were determined by integrating Laboratory results with the clinical diagnoses validated by epileptologists.

As illustrated in the top-right corner of Figure 1A, the initial EHR dataset comprised 40,749 records, including redundant follow-up visits for the patients. After retaining the most recent record for each patient, the dataset was reduced to 5,552 cases. Further exclusions were made for records unrelated to epilepsy or diseases outside the scope of this study, resulting in 590 relevant records. Additional refinement involved the removal of cases with unclear semiology or undetermined EZs. Ultimately, the final dataset consisted of 184 validated semiology-EZ pairs.

Conclusively, we evaluated ChatGPT’s performance using two datasets: a publicly sourced dataset comprising 852 semiology-EZ pairs and a private FEMH dataset containing 184 semiology-EZ pairs.

As illustrated in Figure. 1B-C, the distribution of EZs in both the publicly sourced and private cohorts demonstrates remarkable similarity. In the publicly sourced cohort, the temporal lobe (T) and frontal lobe (F) account for the largest proportions of EZ-semiology cases, followed by the parietal lobe (P) and occipital lobe (O), with the insular cortex (INS) and cingulate cortex (CING) comprising the smallest proportions. Similarly, the private-source cohort exhibits a similar distribution, with a minor variation where the frontal lobe slightly surpasses the temporal lobe in the number of EZ-semiology cases.

The lobe distribution from both private and publicly sourced data cohorts highlights that the temporal and frontal lobes are the most frequently and commonly identified lobes with EZs, not only in the context of research studies but also in real-life clinical practice. Cases with EZs from the parietal and occipital lobes are comparatively less frequent, while those involving the insular cortex and cingulate cortex remain rare, and consistent for private and publicly sourced data cohorts.

### Demographics of patients

The publicly sourced data cohort comprised 852 semiology-EZ pairs from 852 patients. It includes 320 females, 404 males, and 128 patients with undisclosed gender. Of all 852 patients, 134 were right-handed, 22 left-handed, 3 ambidextrous, and 706 had unspecified handedness. The age range of the patients spanned from newborn to 77 years, with 310 individuals under the age of 18 and 335 adults. The average age is 23.66 years old, and the standard deviation is 17.16.

The FEMH data cohort consists of 184 semiology-EZ pairs. This group included 44 female and 46 male patients, and gender information is not available for the others. The age ranges from newborn to 87. Among them, 37 were under 18 years old, and 50 were adults. The average age is 28.67 years old, and the standard deviation is 18.05.

### Response Generation with ChatGPT

For this study, we selected ChatGPT-4 as the primary LLM example due to its superior performance compared to ChatGPT-3.5. The detailed results of ChatGPT-3.5 are included in the appendix for reference. To evaluate the performance of ChatGPT-4, we utilized the “gpt-4-turbo” API and implemented two distinct prompt configurations: zero-shot prompting (ZSP) [21] and few-shot prompting (FSP) [22]

In the ZSP configurations, ChatGPT received no prior examples or guidance and relied solely on its internal knowledge to make predictions. In contrast, the FSP configurations incorporated three randomly selected semiology-EZ pair examples as input, guiding ChatGPT’s predictions to align more closely with the ground truth. These three input examples were held as the same prompts across all test samples to ensure consistency during evaluation.

To standardize query sentence design and ensure robust predictions, all semiology information was formatted into a fixed sentence structure. When patient-specific details were available, the input combined these details with the semiology into a single sentence, such as: “A [handedness] [gender] patient, aged [age], presented with semiology: [semiology].” In cases where patient-specific information was unavailable, the input was simplified to: “The patient presented with semiology: [semiology].” ChatGPT was then tasked with predicting the most likely EZs based solely on the provided semiology, with its output restricted to specifying the EZ location without any accompanying explanation. Examples of query formats and configurations used in this study are provided in Table 1.

**Table 1.** Examples of Questions and Responses Generated by ChatGPT with Different Prompt Configurations.

|  | **Zero-Shot Prompting** | **Few-Shot Prompting** |
| --- | --- | --- |
| **User Input** | One sentence including the patient's semiology and demographic information | One sentence including the patient's semiology and demographic information |
| **Prompt** | None | Case 1:<br>- A right-hand male patient, aged 24, presented with semiology: a slightly painful sensation of tightness in the right shoulder and the region of the right sternocleidomastoid muscle. |
|  |  | - EZ: Frontal lobe<br>Case 2:<br>- A patient presented with semiology: gastric aura and a warm feeling in the chest, rising to her face, facial flushing, followed by oroalimentary automatism<br>- EZ: Temporal lobe<br>Case 3:<br>- A patient presented with semiology: aura, loss of contact, motor.<br>- EZ: Occipital lobe |
| **Query** | Based on the user's semiology, list the most likely epileptogenic zones (EZ) in descending order of likelihood.<br>The epileptogenic zones include frontal lobe, temporal lobe, parietal lobe, occipital lobe, cingulate, insular cortex.<br>Provide the answer in this format: 'EZ1, EZ2, ...'. If there is only one likely EZ, list only that one. Do not include any explanations. | Based on the user's semiology, list the most likely epileptogenic zones (EZ) in descending order of likelihood.<br>The epileptogenic zones include frontal lobe, temporal lobe, parietal lobe, occipital lobe, cingulate, insular cortex.<br>Provide the answer in this format: 'EZ1, EZ2, ...'. If there is only one likely EZ, list only that one. Do not include any explanations. |
| **Description** | A right-handed male patient, aged 38, presented with semiology: speech arrest | A right-handed male patient, aged 38, presented with semiology: speech arrest |
| **Response** | Frontal Lobe | Frontal Lobe, Temporal Lobe |

### Evaluation of seizure semiology interpretation from epileptologists

To evaluate the clinical value of ChatGPT on interpreting seizure semiology, we assessed its performance by comparing it to a panel of Board-certified epileptologists [23] using the voluntary seizure semiology survey [24]. The research team initially tested the survey’s usability and technical functionality before its public debut. After optimization and improvement, the final version was deployed for response collection. This survey was conducted using an open online survey platform named Zoho Survey, which is a free online tool that facilitates automated survey creation and response collection. It was structured into multiple sections to ensure clarity and ease of participation. The first page provided participants with essential instructions, including the total number of questions, the estimated time required, guidelines on completing each query, and instructions on how to save progress. An example was also included for better instruction. The second page collected bibliographic details of participants, which were strictly protected and used solely for research purposes. From the third page onward, participants were required to complete 100 compulsory multiple-choice questions, each presenting a unique seizure semiology description. Participants were asked to determine the most likely EZ from six options: frontal lobe, temporal lobe, parietal lobe, occipital lobe, cingulate cortex, and insular cortex, with an additional “Other” comment box for alternative responses. Each page displayed a single patient case, and participants could proceed to the next question by clicking the “Next” button. If a participant wished to revise a previous response, they could navigate back using the “Previous” button. The last page demonstrates appreciation for participants.

To track and analyze the participant’s performance, the Zoho platform automatically recorded visitor IP addresses to determine the total number of unique participants and tracked survey progress, allowing researchers to identify whether a participant only completed the bibliographic section or proceeded to answer the questions. To prevent data loss, participants were instructed to press the “Save and Continue Later” button (available from the second page onward) if they needed to take a break. Additionally, surveys submitted with atypical timestamps were flagged for further review to ensure data integrity and reliability.

To minimize selection bias regarding region or semiology, we randomly selected a subset of 100 semiology records from our self-compiled database, covering all six general brain regions. The final subset used in the survey met the following criteria: (1) selected semiology records contained comprehensive and explicit descriptions of seizure symptoms; (2) the distribution of EZs spanned all six general regions, rather than focusing on one region; and (3) the records were chosen to capture the widest possible range of seizure symptoms.

Epileptologists were recruited through the National Association of Epilepsy Center and the American Epilepsy Society, with over 70 survey invitation emails with survey links distributed worldwide. Responses were collected between January 2024 and July 2024. A total of eight epileptologists participated, of whom six completed the survey in full, while two provided partial responses. All participating epileptologists were employed at different epilepsy centers during the survey period, with clinical experience ranging from 7 to 35 years. Their affiliations spanned across the western, central, and eastern regions of the United States, as well as Germany, ensuring geographic diversity.

For comparative analysis, five epileptologists were selected from the six who completed the survey given one epileptologist was primarily employed as a neuro-oncologist. These selected specialists served as a benchmark for evaluating ChatGPT’s accuracy in EZ-based semiology prediction. To ensure consistency, the input format presented to the epileptologists mirrored the structure of the queries used for ChatGPT. Epileptologists were tasked with identifying the most likely EZ locations based on the provided semiology. Comparative examples of outputs from ChatGPT and the selected epileptologists are presented in Table 2.

**Table 2.** Performance Comparison of ZSP Semiology Predictions Among GPT-4 and E1–E5.

| No. | Semiology Interpretation Query | Ground Truth | GPT-4 | E1 | E2 | E3 | E4 | E5 |
| --- | --- | --- | --- | --- | --- | --- | --- | --- |
| 1 | The patient is a 14-year-old right-hand male. His reported semiology is speech arrest and lower speech intensity.<br>According to the patient's reported semiology, what are the most likely general lobes of the brain where the epileptogenic zone is located? (multiple choices question) | INS | F, INS | F | F | F, T | F, INS | F, T |
| 2 | The patient, a right-handed 67-year-old female, began experiencing seizures at the age of 66. During 2 of the 9 recorded seizures, she exhibited symptoms of drooling, spitting, and coughing.<br>According to the patient's reported semiology, what are the most likely general lobes of the brain where the epileptogenic zone is located? (multiple choices question) | P | T, INS | T, INS | T | F, INS | F, INS | F |
| 3 | The patient is a 43-year-old left-hand male. His age at seizure onset was 6 years old. His reported semiology is a dreamy state, loss of consciousness, oroalimentary and gestural automatisms, nose wiping, and right arm dystonic posturing.<br>According to the patient's reported semiology, what are the most likely general lobes of the brain where the epileptogenic zone is located? (multiple choices question) | T | T | T | T | F, T | T | T |

### Statistical Analysis

The inference of EZ location is determined using the six-lobe classification criteria. To evaluate the responses from ChatGPT and epileptologists, we used three metrics: Regional Sensitivity (RSens), Weighted Sensitivity (WSens), and Net Positive Inference Rate (NPIR).

Sensitivity is a widely used metric in classification problems [25]. In this study, we not only calculate the overall sensitivity but also evaluate the sensitivity for each EZ region. Specifically, RSens measured the accuracy of ChatGPT or epileptologists in identifying the correct region and is defined as follows:

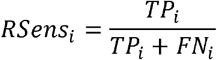

where *i* denotes the index corresponding to six general regions, *Rsen*_*i*_ denotes the sensitivity value for region *i*,*TP*_*i*_ (True Positive) denotes the number of correctly identified EZs, and *FN*_*i*_ (False Negative) denotes the number of EZs that were not correctly identified.

For example, consider a dataset of 100 semiology-EZ samples. The region mapping is as follows: 0-Cingulate (CING), 1-Frontal (F), 2-Insular (INS), 3-Occipital (O), 4-Parietal (P), and 5-Temporal (T). If 80 of these cases have the true label F, but only 60 are correctly identified as F (*TP*_1_ = 60), while the remaining 20 are mislabeled as other regions (*FN*_*i*_ = 20), the sensitivity for the frontal lobe is calculated as: 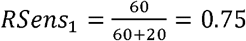. This means that 75% of the F cases are correctly identified.

Additionally, given the unbalanced distribution of EZs across the six general regions, we addressed the class imbalance issue by using WSens to provide a more accurate performance assessment. WSens evaluates overall accuracy by considering the RSens of each region and its corresponding weight in the dataset, which is calculated as follows:

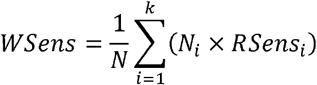

where *k* denotes the total number of regions, *i* denotes the index corresponding to each region; *N* denotes the total number of regions in the dataset, and *N*_*i*_ denotes the count of instances for the *i*-th region.

For instance, consider a dataset with 130 cases distributed across six regions, with the distribution of instances (*N*_*i*_) and their corresponding *Rsen*_*i*_ as follows: *N*_*0*_ = 10, *Rsen*_0_ = 0.1; *N*_1_ = 40, *Rsen*_1_ = 0.7; *N*_2_ = 5, *Rsen*_2_ = 0.5, *N*_2_ = 15, *Rsen*_3_ = 0.6; *N*_4_ = 20, *Rsen*_4_ = 0.4; and *N*_5_ = 40, *Rsen*_2_ = 0.58. The N is calculated as: *N* = 10+ 40+5 + 15 + 20+ 40+ 130, and the WSens is then computed as: 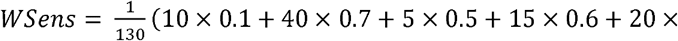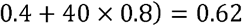. This indicates an overall accuracy of 62% across six regions.

While sensitivity for an individual region and across six regions have been extensively discussed, the evaluation of performance for each semiology query – representing an individual patient’s case – has not been addressed. This aspect is particularly significant for clinical practice. To bridge this gap, the NPIR (net positive inference rate) is introduced, derived from RSens. Although the denominator remains unchanged, the numerator is adjusted to reward correctly inferred regions for a given semiology while penalizing incorrect inference. The NPIR is calculated as follows:

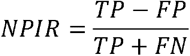

where *TP* (True Positive) is the number of correctly identified regions, *FP* (False Positive) is the number of regions incorrectly identified, and *FN* (False Negative) is the number of regions that were part of the ground truth but not identified. For example, consider a patient’s semiology query with the true labels F and T. If the prediction includes T, P, and O, then *TP* = 1 (for T, correctly identified), *FP* = 2 (for P and O, incorrectly identified), and *FP* = 1 (for F, missed in the prediction). The NPIR is: 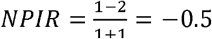.This result indicates that the model provides partially correct predictions but also identifies misleading EZ locations.

Regarding value interpretation, an NPIR value reflects the reliability of the inference results. An NPIR of 1 indicates a completely correct inference of the EZ location. A value below 1 suggests the inference is partially incorrect or contains omissions. An NPIR below 0 indicates that the inference is unreliable and could mislead physicians during preoperative assessments for epilepsy surgery.

In summary, RSens quantifies the ability to accurately identify EZs for specific brain regions and provides insights into a tool’s performance on individual regions in a clinical context. Although RSens values range between 0 and 1, they are better interpreted as percentages to represent the proportion of correctly identified EZs in a given region. Similarly, WSens accounts for class imbalance by weighting regional sensitivities, offering an overall accuracy metric that reflects performance across all regions. Like RSens, WSens Should also be interpreted as a percentage for ease of understanding in clinical applications.

By contrast, NPIR assesses the reliability of predictions by penalizing incorrect inferences. Higher NPIR scores indicate more trustworthy results, while negative scores highlight misleading predictions, both of which are critical for clinical practice. However, NPIR is influenced by the number of predicted regions provided by ChatGPT or epileptologists. When multiple EZ predictions are made for a single case, NPIR values often cluster around 0 or fall below 0, reflecting the trade-off between partially correct and incorrect inferences.

## Results

### Evaluation of Responses from ChatGPT on Publicly Sourced Cohort

In this section, we evaluated the performance of ChatGPT-4 using ZSP (abbreviated as GPT-4 ZSP), and ChatGPT-4 using FSP (GPT-4 FSP) in interpreting seizure semiology based on the publicly sourced cohort. The evaluation results for ChatGPT-4 are presented in Figure 2, while those for ChatGPT-3.5 are provided in Appendix.

**Figure 2:**
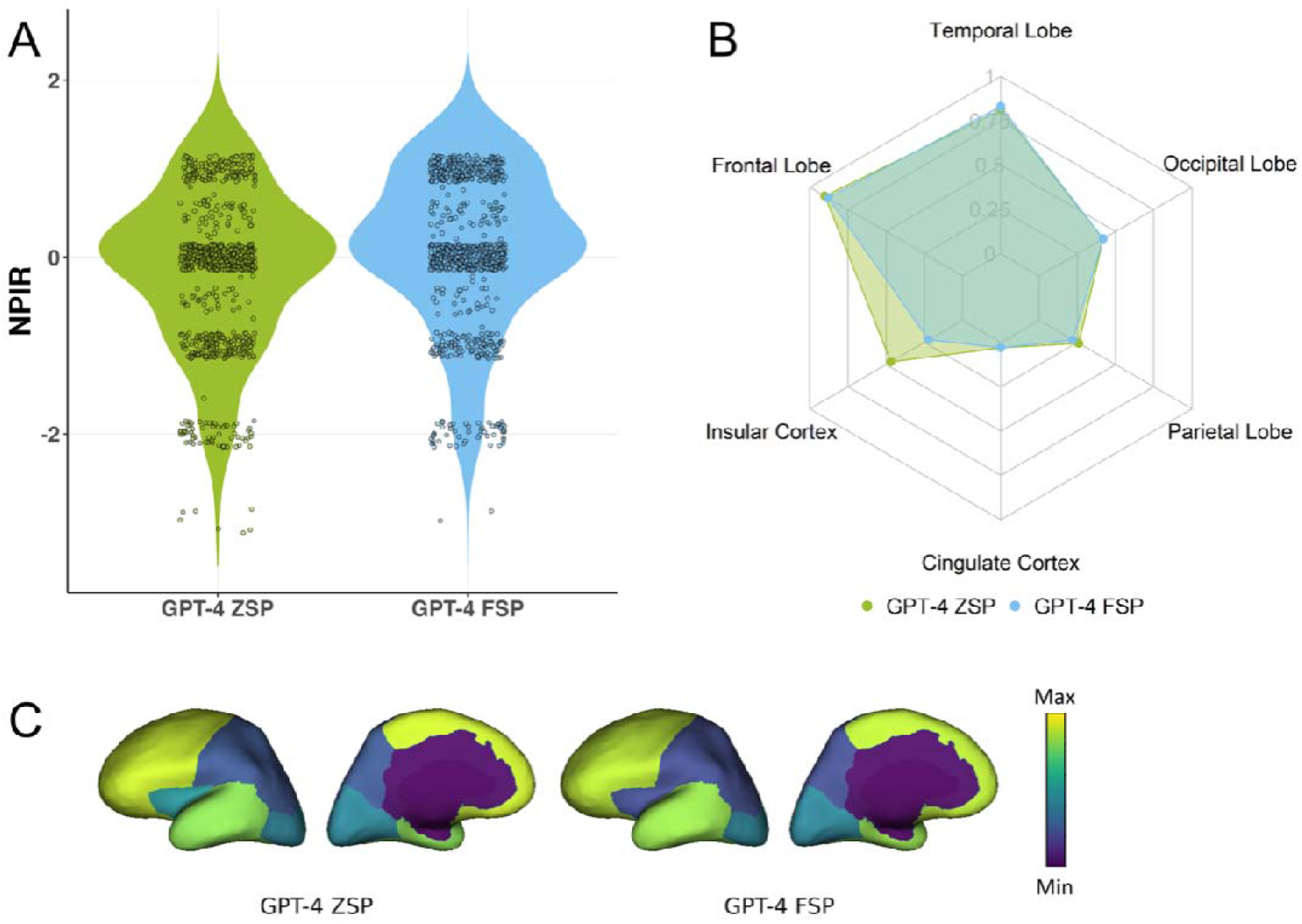
(A) Net Positive Inference Rate Distribution generated by ChatGPT-4 with zero-shot prompting (GPT-4 ZSP) and few-shot prompting (GPT-4 FSP). (B) and (C), regional sensitivity generated by ChatGPT with different prompting configurations (GPT-4 ZSP, GPT-4 FSP).

The RSens values for each region were presented in Figure 2B-C. For the frontal lobe, GPT-4 ZSP achieved 0.90 and FSP achieved 0.88; for the temporal lobe, ZSP achieved 0.81 and FSP achieved 0.83; for the occipital lobe, both ZSP and FSP achieved 0.42; for the parietal lobe, ZSP achieved 0.26 and FSP achieved 0.22; for the insular cortex, ZSP achieved 0.47 and FSP achieved 0.22; and for the cingulate cortex, both ZSP and FSP achieved 0.03.

These RSens results highlight ChatGPT’s proficiency in interpreting seizure semiology for the frontal and temporal lobes, which are the most commonly observed regions in both the public and private data cohorts. However, performance declines for the occipital lobe, parietal lobe, and insular cortex, which are less frequently represented in the distribution. For the cingulate cortex, the least commonly involved region as observed in both data cohorts, performance remained consistently low across both prompting methods.

In terms of WSens, both GPT-4 ZSP and GPT-4 FSP demonstrated comparable results, with WSens values of 0.69 and 0.67, respectively. Although these values are not exceptionally high, they should be evaluated in comparison to the performance of epileptologists to determine the relative accuracy and potential utility of ChatGPT-4 in assisting seizure semiology interpretation.

As shown in Figure. 2A, the NPIR values, which reflect the interpretation accuracy for each semiology prediction, indicate that GPT-4 ZSP achieved a mean of -0.21, while GPT-4 FSP demonstrated a slightly higher mean of 0.03. An NPIR close to 0 indicates that ChatGPT-4’s predictions often include at least one correctly identified region but may also involve one misleading region.

### Evaluation of Responses from ChatGPT with Private-Source Cohort

Given that all papers used to compile the publicly sourced database are available online, some may have been included in the training corpus of ChatGPT, potentially making the evaluation results less objective and convincing. To address this concern, we employed a database with a private source for external validation of ChatGPT’s performance. The evaluation results for this database are presented in Figure. 3.

**Figure 3:**
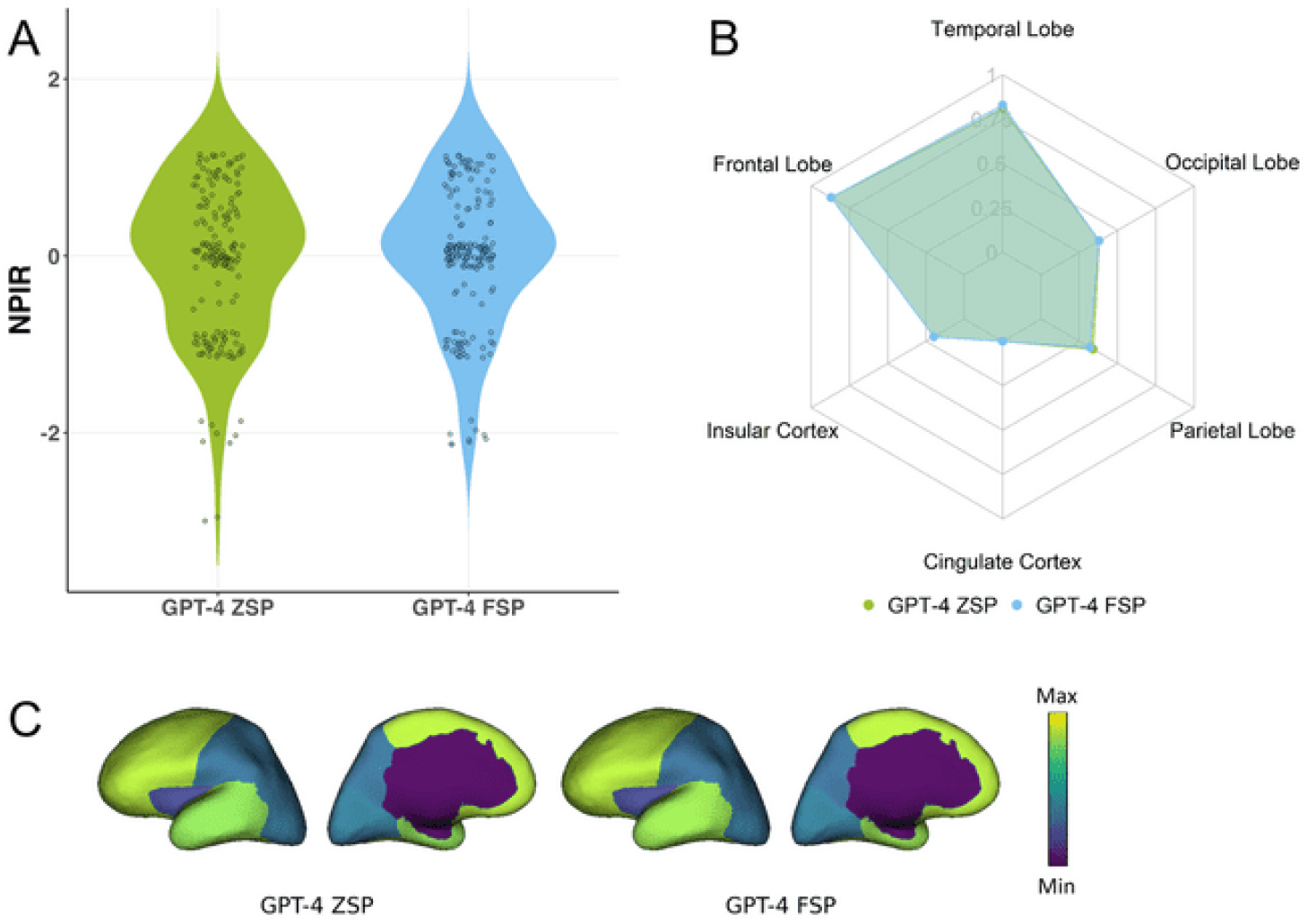
(A) Net Positive Inference Rate Distribution generated by ChatGPT-4 with zero-shot prompting (GPT-4 ZSP) and few-shot prompting (GPT-4 FSP). (B) and (C), regional sensitivity generated by ChatGPT with different prompt configurations (GPT-4 ZSP, GPT-4 FSP).

The RSens values for each brain region are presented in Figure. 3B-C. For the frontal lobe, both GPT-4 ZSP and FSP achieved an RSens value of 0.87. For the temporal lobe, ZSP achieved 0.81, while FSP achieved 0.83. For the occipital lobe, both methods achieved 0.38, and for the parietal lobe, ZSP achieved 0.34, while FSP achieved 0.32. For the insular cortex, both ZSP and FSP achieved 0.20, and for the cingulate cortex, both recorded a value of 0.

In terms of WSens, GPT-4 ZSP achieved a value of 0.73, while FSP slightly outperformed it with 0.74. For NPIR, GPT-4 ZSP achieved a mean of -0.20, while FSP demonstrated a slightly improved mean of -0.12, as shown in Figure. 3A.

These evaluation results based on the private-source cohort are consistent with those from the publicly sourced cohort, further confirming ChatGPT’s ability to interpret seizure semiology across different brain regions.

### Comparison of Responses from ChatGPT and Epileptologists

In this section, we compared the performance of ChatGPT with that of 5 board-certified epileptologists (EP-1, EP-2, EP-3, EP-4, EP-5). The comparison results are shown in Figure. 4.

**Figure 4:**
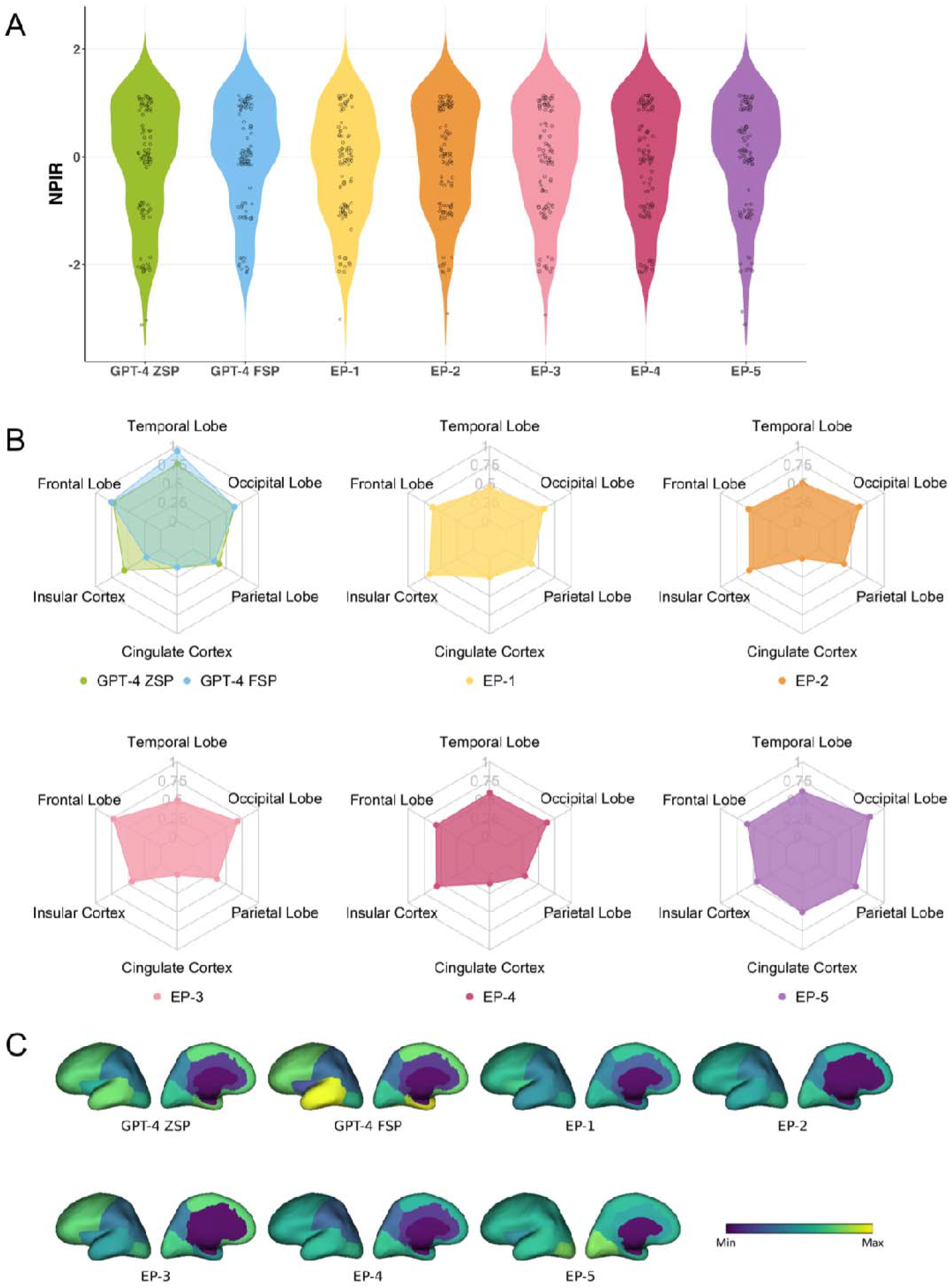
(A) Net Positive Inference Rate Distribution generated by ChatGPT-4 with zero-shot prompting (GPT-4 ZSP) and few-shot prompting (GPT-4 FSP). (B) and (C), regional sensitivity generated by ChatGPT-4 with different prompt configurations (GPT-4 ZSP, GPT-4 FSP).

When comparing RSens values across regions (Figure. 4B-C), ChatGPT-4 *outperformed* epileptologists in interpreting seizure semiology for the frontal (ZSP: 0.73, FSP: 0.76, EPs: 0.57–0.73) and temporal lobes (ZSP: 0.76, FSP: 0.93, EPs: 0.44–0.61), and demonstrated comparable performance for the parietal (ZSP: 0.39, FSP: 0.32, EPs: 0.29–0.57), occipital lobes (ZSP: 0.63, FSP: 0.63, EPs: 0.58–0.79), and insular cortex (ZSP: 0.56, FSP: 0.22, EPs: 0.44–0.67). However, it *underperformed* epileptologists in interpreting seizure semiology cases that are associated with the cingulate cortex (ZSP/FSP: 0.12, EPs: 0–0.5). These results indicate that ChatGPT-4 outperforms epileptologists in interpreting seizure semiology for regions that are more commonly represented, such as the frontal and temporal lobes. However, its performance declines for less frequently observed regions in both the public databases (representing research settings) and private databases (reflecting real-world clinical contexts).

In terms of WSens, ChatGPT-4 significantly outperformed the epileptologists. Specifically, GPT-4 ZSP achieved a WSens of 0.61, and GPT-4 FSP achieved a slightly higher WSens of 0.63. In comparison, the WSens of epileptologists ranged from 0.49 (EP-2) to 0.51 (EP-5). These findings highlight that ChatGPT-4 delivers more consistent accuracy across all regions compared to epileptologists.

For NPIR, as illustrated in Figure. 4A, GPT-4 ZSP achieved a mean of -0.14, while GPT-4 FSP achieved a mean of -0.02. Among the epileptologists, EP-5 achieved the highest mean NPIR of -0.08, while EP-2 recorded the lowest mean of - 0.13. The performance of both GPT-4 and the epileptologists remained near 0, indicating comparable performance between the two.

### Significant Testing of Responses from ChatGPT and Epileptologists

To evaluate whether the differences in RSens performance between ChatGPT and epileptologists are statistically significant, we utilized data from the survey of 100 semiology cases. A group t-test was conducted with 1000 bootstrap samples to determine the significance of these differences. The comparison focused on the averaged RSens performance of ChatGPT-4 (using ZSP and FSP) and the averaged performance of five epileptologists. As illustrated in Figure. 5A, the results indicated significant differences, suggesting that ChatGPT-4 excels in commonly represented EZs like the frontal lobe and temporal lobe but faces challenges in rarer EZs, such as the cingulate cortex.

**Figure 5:**
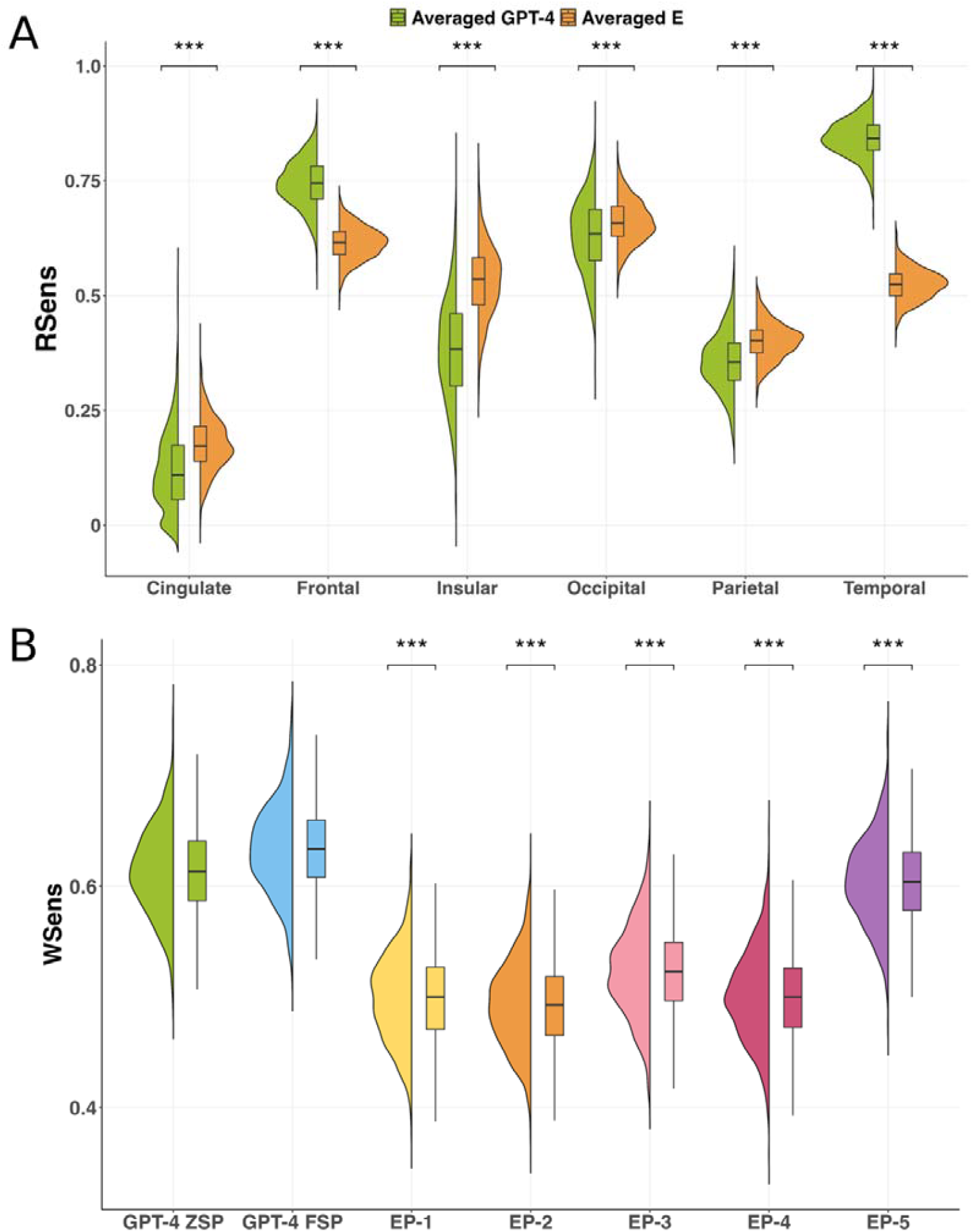
(A) Significance Testing of Regional Sensitivity (RSens) comparing Averaged GPT-4 (Zero-Shot and Few-Shot Prompting) with Averaged Epileptologists (Averaged E). (B) Significance Testing of Weighted Sensitivity (WSens) for GPT-4 (Zero-Shot and Few-Shot Prompting) and Five Individual Epileptologists (EP-1 to EP-5).

For WSens, a similar group t-test with 1000 bootstrap samples was applied to assess the overall performance differences between ChatGPT-4 using ZSP, ChatGPT-4 using FSP, and the five epileptologists. As shown in Figure. 5B, ChatGPT-4 statistically significantly outperformed the group of epileptologists in overall sensitivity. Among the five epileptologists, E1 and E4 did not show a statistically significant difference in performance, whereas the remaining epileptologists demonstrated significant differences from one another, with p-values less than 0.001.

Interestingly, the years of clinical experience among epileptologists, which ranged from 7 to 35 years, did not show a consistent correlation with performance. For instance, E1 (20 years of experience) and E4 (8 years of experience) demonstrated similar performance levels, while E3, the least experienced with 7 years of practice, ranked among the top two performers. In contrast, E2 (16 years of experience) showed the lowest performance, whereas E5, the most experienced with 35 years of practice, achieved the highest performance in the group. These findings suggest that clinical experience alone may not guarantee consistent reliability, highlighting the potential value of supplementary tools, such as ChatGPT, to assist in clinical decision-making.

For NPIR, as shown in Section 3.3 and **Figure. 5A**, the distributions of NPIR values for ChatGPT-4 ZSP, ChatGPT-4 FSP, and the five epileptologists are visually similar. Given we already have data distributions, a direct t-test was conducted on the 100 survey entries. The results indicate no statistically significant differences in NPIR performance between ChatGPT-4 and the epileptologists.

## Discussion

### Main Findings

We evaluated the capability of ChatGPT on interpreting seizure semiology to localize the EZ using a publicly sourced cohort with 852 Semiology-EZ pairs from peer-reviewed journals, a private-source cohort with 184 pairs from FEHM hospital, and a randomly selected 100 semiology survey.

For the analysis of RSens, ChatGPT achieved RSens values of approximately 80-90% in the most commonly observed regions, such as the frontal and temporal lobes, in both the public and private databases. However, RSens declined as the regions became less common, including the occipital lobe, parietal lobe, and insular cortex, and dropped significantly for rare regions like the cingulate cortex, where ChatGPT failed to predict correctly in most cases. These findings align with the survey comparison between ChatGPT and epileptologists, where ChatGPT significantly outperformed epileptologists in identifying EZ locations in more common regions, demonstrated comparable but slightly lower performance in less common regions, and substantially underperformed in rare regions.

The higher performance on common EZ-based semiology interpretation query and lower performance on less common questions observed in this study is consistent with findings from previous studies assessing ChatGPT’s performance in epilepsy-related inquiries. Specifically, Kim et al. assessed the reliability of responses of ChatGPT to 57 commonly asked epilepsy questions, and all responses were reviewed by two epileptologists. The results suggested that these responses were either of “sufficient educational value” or “correct but inadequate” for almost all questions [26]. Wu et al. evaluated the performance of ChatGPT to a total of 378 questions related to epilepsy and 5 questions related to emotional support. Statistics indicated that ChatGPT provided “correct and comprehensive” answers to 68.4% of the questions. However, when answering “prognostic questions”, ChatGPT performed poorly with only 46.8% of answers rated comprehensive [27]. These findings suggest that ChatGPT’s performance is positively correlated with the availability of sufficient data to support its responses.

Although ChatGPT’s performance on RSens varies depending on the EZ localization of seizure semiology, it remains clinically useful because, in most real-world cases, patient symptoms align with more common seizure types, such as frontal lobe and temporal lobe seizures. These two types account for 80% of cases in the private data cohort from the real-life hospital. Furthermore, for WSens, ChatGPT achieved over 60% accuracy in both public and private databases and significantly outperformed five epileptologists, demonstrating its strong generalizability in seizure semiology interpretation.

Given these findings, ChatGPT could serve different roles depending on the clinical setting. In epilepsy centers with rich resources, it may function as a co-pilot to support epileptologists in improving diagnostic efficiency. In resource-limited epilepsy centers, where access to specialized epilepsy care is scarce, ChatGPT could be particularly valuable in assisting general practitioners or non-specialist clinicians with preliminary seizure classification and decision-making, potentially improving access to epilepsy care.

Besides those metrics, ChatGPT has the potential to further assist and enhance clinical decision-making in epilepsy centers to achieve optimal post-surgical outcomes since it could make pre-surgical EZ predictions based on an LLM trained with a large cohort of cases, while epileptologists could only predict through their clinical experience, laboratory results and patients’ self-report.

## Limitations

Although this study offered an important reference on the capability of ChatGPT to interpret the descriptions of seizure semiology to localize EZs, there are still several limitations.

Firstly, when inferring the EZ location according to semiology, the identified area is referred to as the Symptomatogenic Zone (SZ), which is the region responsible for the observed seizure symptoms but may not fully align with the actual EZ which is a theoretical definition given by Dr. Hans Lüders [8] and whose removal will make the patients seizure-free thus can only be derived after surgical oblation. This will result in limited precision in predicting EZs. Additionally, epileptic seizures often involve abnormal activities across multiple brain regions, with certain symptoms arising from the propagation of activity to regions beyond the EZ, which may lead to potential misjudgments.

Secondly, the database used to train and evaluate ChatGPT in this study could be further expanded and refined. The two databases both have inadequate data for less common regions such as the cingulate cortex and insular cortex. Additionally, ChatGPT was trained on the Common Crawl corpus, which encompasses a broad range of general knowledge but lacks a specific focus on medical or epilepsy-related information. This lack of specialized data limits ChatGPT’s ability to generate precise and reliable responses for semiology interpretation and EZ localization.

Thirdly, there may be biases in ChatGPT’s output due to the underlying training cohorts. For instance, the datasets predominantly include cases from Europe, East Asia, and North America, with significantly fewer cases from South America and Africa. Additionally, the data largely represents patients who can afford epilepsy surgery. As a result, predictions might be influenced by this non-representative sampling of patients from around the world.

Lastly, the limited number of participating epileptologists inherently restricts the sample size for more comprehensive analysis. Insights from a small group of epileptologists may not adequately reflect the broader expertise and perspectives of epileptologists from the global community.

## Future Work

As mentioned in the study, there is an imbalance in semiology data distribution across the six lobes, with most cases originating from the frontal and temporal lobes. To mitigate this bias in the current study, we have incorporated weighted sensitivity analysis to account for differences in EZ distribution. Additionally, we plan to address this limitation in future studies by collecting more data on less common epilepsy regions and fine-tuning LLMs using epilepsy-specific corpora. In addition, a sequential description of seizure semiology can help map out the seizure propagation pathways from seizure onset zones to other symptomatogenic brain regions, it is important to leverage the sequence information of seizure semiology to provide a detailed characterization of the epileptic brain network.

## Acknowledgments

The authors are grateful to the epileptologists who completed the survey.

## Data Availability

The data will be made available upon reasonable request to the corresponding author Dr. Feng Liu.

## Author Contributions

Jiao (data curation, formal analysis, software, Visualization, Writing - Original Draft), Luo (data curation, formal analysis, software, Visualization,Writing - Original Draft), Fotedar (Conceptualization, Methodology, Validation, Investigation, Writing - Review & Editing), Karakis (Validation, Investigation, Writing - Review & Editing), Aboud (Validation, Investigation, Writing - Review & Editing), Rao (Validation, Investigation, Writing - Review & Editing), Asmar (Validation, Investigation, Writing - Review & Editing), Xian (Statistical Analysis, Validation, Writing - Review & Editing), Wen(Statistical Analysis, Validation, Writing - Review & Editing), Ding (Validation, Writing - Review & Editing), Lin (Resources, Validation, Writing - Review & Editing), Rosenow (Validation, Investigation, Writing - Review & Editing), Sun (Conceptualization, Methodology, Validation, Investigation, Writing - Review & Editing), Liu (Conceptualization, Supervision, Formal analysis, Funding acquisition, Resources, Methodology, Writing - Original Draft)

## Abbreviations

ChatGPT: Chat Generative Pre-trained Transformer
CING: Cingulate Cortex
EHR: electronic health record
EP-1: Epileptologist 1
EP-2: Epileptologist 2
EP-3: Epileptologist 3
EP-4: Epileptologist 4
EP-5: Epileptologist 5
EZ: Epileptogenic Zone
F: Frontal Lobe
FEMH: Far Eastern Memorial Hospital
GPT: Generative Pre-trained Transformer
INS: Insular Cortex
LLM: large language model
NPIR: Net Positive Inference Reliability
O: Occipital lobe
P: Parietal Lobe
RSens: Regional Sensitivity
T: Temporal Lobe
WSens: Weighted Sensitivity

## Multimedia Appendix 1

(A) Net Positive Inference Rate (NPIR) distribution for GPT-3.5 with zero-shot prompting (ZSP) and few-shot prompting (FSP) on the publicly sourced database. (B) Regional sensitivity (RSens) for GPT-3.5 ZSP and FSP on the publicly sourced database. (C) NPIR distribution on the private-source database. (D) RSens on the private-source database. (E) NPIR distribution on the 100-question survey. (F) RSens on the 100-question survey.

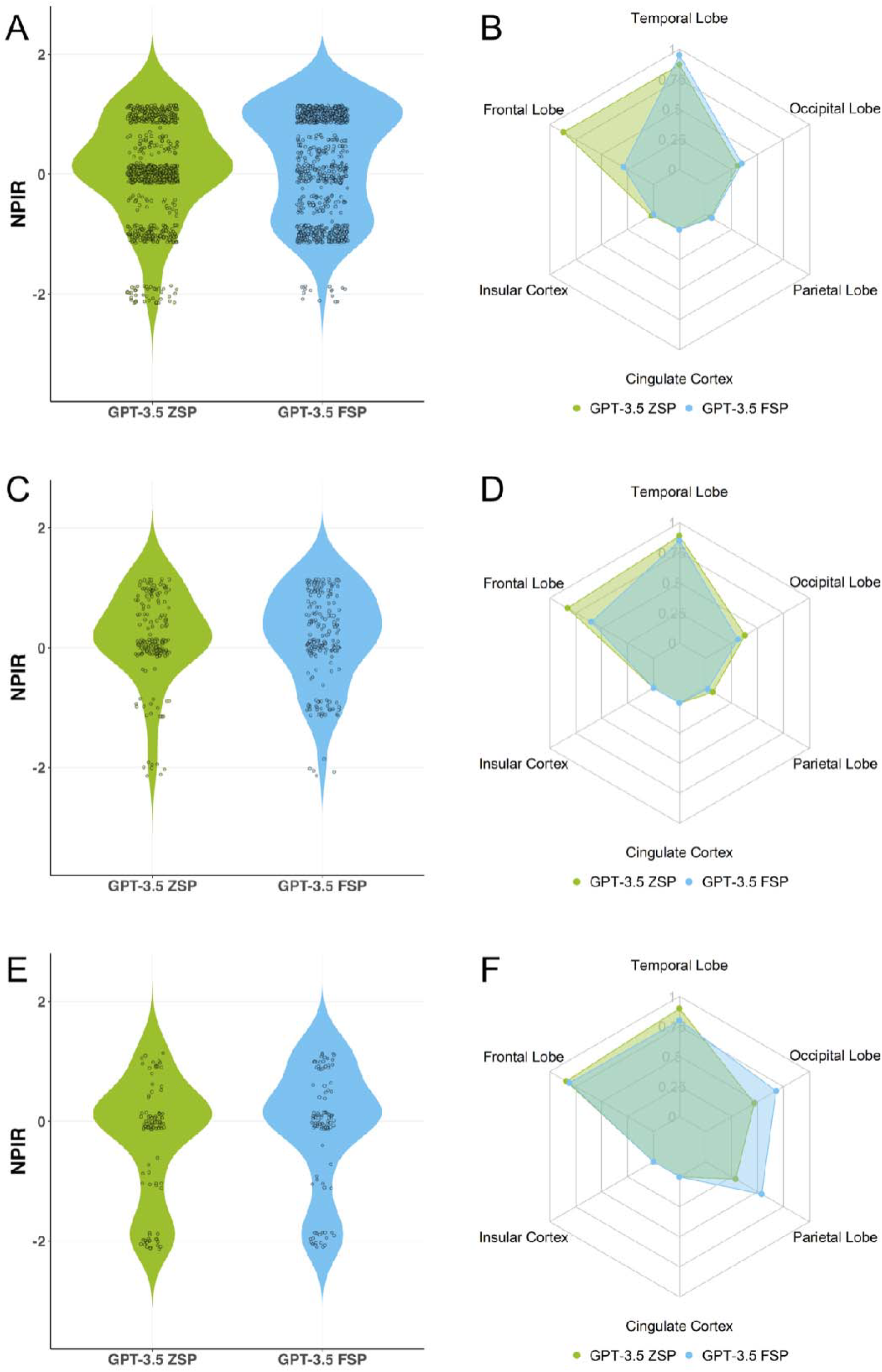

